# Understanding microvascular thrombosis in COVID-19 via massive single-cell imaging of circulating platelets

**DOI:** 10.1101/2021.04.29.21256354

**Authors:** Hiroshi Kanno, Yuqi Zhou, Masako Nishikawa, Ting-Hui Xiao, Takuma Suzuki, Yuma Ibayashi, Jeffrey Harmon, Shigekazu Takizawa, Kotaro Hiramatsu, Nao Nitta, Risako Kameyama, Walker Peterson, Jun Takiguchi, Mohammad Shifat-E-Rabbi, Yan Zhuang, Xuwang Yin, Abu Hasnat Mohammad Rubaiyat, Yunjie Deng, Hongqian Zhang, Shigeki Miyata, Gustavo K. Rohde, Wataru Iwasaki, Yutaka Yatomi, Keisuke Goda

**Affiliations:** Department of Chemistry, The University of Tokyo, Tokyo 113-0033, Japan; Department of Clinical Laboratory Medicine, Graduate School of Medicine, The University of Tokyo, Tokyo 113-0033, Japan; Department of Computational Biology and Medical Sciences, The University of Tokyo, 277-8562, Japan; Research Center for Spectrochemistry, The University of Tokyo, Tokyo 113-0033, Japan; CYBO, Inc., Tokyo 101-0022, Japan; Department of Biomedical Engineering, University of Virginia, Virginia 22908, USA; Department of Electrical and Computer Engineering, University of Virginia, Virginia 22908, USA; Research and Development Department, Central Blood Institute, Japanese Red Cross Society, Tokyo 135-8521, Japan; Department of Biological Sciences, The University of Tokyo, Tokyo 113-0033, Japan; Department of Integrated Biosciences, The University of Tokyo, Tokyo 277-8562, Japan; Institute of Technological Sciences, Wuhan University, 430072 Hubei, China; Department of Bioengineering, University of California, Los Angeles, California 90095, USA

## Abstract

A characteristic clinical feature of COVID-19 is the frequent incidence of microvascular thrombosis. In fact, COVID-19 autopsy reports have shown widespread thrombotic microangiopathy characterized by extensive diffuse microthrombi within peripheral capillaries and arterioles in lungs, hearts, and other organs, resulting in multiorgan failure. However, the underlying process of COVID-19-associated microvascular thrombosis remains elusive due to the lack of tools to statistically examine platelet aggregation (i.e., the initiation of microthrombus formation) in detail. Here we present a method for massive image-based profiling, temporal monitoring, and big data analysis of circulating platelets and platelet aggregates in the blood of COVID-19 patients at single-cell resolution, to provide previously unattainable insights into the disease. In fact, our analysis of the image data from 110 hospitalized patients shows the anomalous presence of excessive platelet aggregates in nearly 90% of all COVID-19 patients. Furthermore, results indicate strong links between the concentration of platelet aggregates and the severity, mortality, and respiratory condition of patients with COVID-19. Finally, high-dimensional analysis based on deep learning shows that the disease behaves as systemic thrombosis.

## MAIN

With the increasing number of global case reports since the beginning of the COVID-19 pandemic, it has become clear that COVID-19-associated microvascular thrombosis is one of the key factors for the severity and mortality of COVID-19^1,2,3,4,5,6,7,8,9,10,11^. In fact, earlier autopsy reports on patients who died with COVID-19 have shown widespread thrombotic microangiopathy (TMA) characterized by extensive diffuse microthrombi present within peripheral capillaries and arterioles in lungs, hearts, and other organs, resulting in multiorgan failure^1,3,7,8,9,10,11^. This is aligned with respiratory failure due to severe diffuse alveolar damage being the primary cause of death in COVID-19^3,10^. It is also consistent with our current understanding of the pathophysiological mechanism of COVID-19 in which SARS-CoV-2’s entry into host cells is mediated by the angiotensin-converting enzyme 2 (ACE2) receptor, which stimulates the renin-angiotensin-aldosterone system and causes vascular endothelial damage followed by vasculitis (i.e., the inflammation of blood vessels), resulting in the formation of microthrombi^2,6^. In response to a number of reports that anticoagulant therapy with heparin leads to better prognosis in COVID-19 patients^12^, both domestic and international organizations have issued clinical practice guidelines recommending that all hospitalized COVID-19 patients should receive thromboprophylaxis (mainly heparin treatment) even without clear symptoms of thrombotic complications and understanding their efficacy^13,14^.

Unfortunately, the underlying process of the incidence of COVID-19-associated microvascular thrombosis remains elusive. This is due to the lack of tools to statistically examine the detailed characteristics of platelet activity, or more specifically platelet aggregation (i.e., the initiation of thrombus formation)^15,16,17,18^ in vivo. For example, optical microscopy^19^ can directly probe platelet aggregation with high spatial resolution and has, in fact, visualized platelet hyperactivity in COVID-19 by identifying the presence of platelet aggregates (i.e., thrombus constituents) including macrothrombocytes^20^. However, visual inspection under the optical microscope is too slow and labor-intensive to analyze platelet aggregates in a statistically meaningful manner. Flow cytometry, on the other hand, enables statistical analysis of large populations of cells by measuring their physical and chemical properties via impedance, scattering, or fluorescence measurements in flow^21,22,23,24,25^ and has been used to identify leukocyte hyperactivity in COVID-19 via fluorescent probes^26,27,28^. However, the lack of spatial resolution in flow cytometry prohibits accurate differentiation between single platelets and platelet aggregates^24,29,30^ and hence cannot accurately characterize platelet activity. Likewise, light transmission aggregometry^31^, although widely used in clinical laboratory testing, only evaluates the average agonist-induced aggregation properties of many platelets in vitro through ensemble measurements and cannot provide details of platelet aggregation such as the size distribution or single-to-aggregate population ratio. Moreover, D-dimer testing, another widely used diagnostic, is conventionally used to estimate the presence of thrombi by measuring the cross-linked fibrin monomers (called D-dimers) produced when thrombi are degraded by fibrinolysis^32^. D-dimer testing has been reported as useful for assessing the severity of COVID-19, as an elevated level of D-dimers (>1 µg/mL) at admission is associated with an increased risk of both required mechanical ventilation and death due to complications^1,3,9,12,33,34^. However, D-dimer testing does not probe platelet activity in vivo and is not sensitive to microvascular thrombosis including TMA (unless it becomes severe)^35^. Medical imaging techniques such as computed tomography (CT), magnetic resonance imaging (MRI), and ultrasonography can directly identify thrombi in the body, but do not have sufficient spatial resolution to recognize circulating platelet aggregates and microthrombi^36^. Finally, postmortem examination can directly identify microthrombi but is only applicable to dead patients. To date, statistical morphometric understanding of platelet aggregation has been inaccessible and hence overlooked, as optical microscopy (a high-content but low-throughput tool) has been the main method used to examine platelet aggregation in detail thus far^24,30^. A new tool is clearly needed to probe the transient process of microthrombus formation and shed light on the pathophysiological mechanism of COVID-19-associated microvascular thrombosis.

In this Article, to overcome the above technical limitations and better comprehend microthrombus formation in COVID-19, we present a method for massive image-based profiling, temporal monitoring, and big data analysis of circulating platelets and platelet aggregates in the blood of COVID-19 patients at single-cell resolution. As shown in Figure 1a, this method consists of (i) blood draw (1 mL) from COVID-19 patients (Supplementary Table 1); (ii) sample preparation by isolating platelets and platelet aggregates from the blood; (iii) high-throughput, blur-free, bright-field imaging of a large population (n = 25,000) of single platelets and platelet aggregates (including platelet-platelet aggregates and platelet-leukocyte aggregates^37^) in blood samples, by optical frequency-division-multiplexed (FDM) microscopy^30^ on a hydrodynamic-focusing microfluidic chip^38,39^ (Figure 1b, Figure 1c, Supplementary Figure 1, see “optical frequency-division-multiplexed microscope” in the Methods section for details); and (iv) digital image processing and the application of various techniques for statistical platelet aggregate analysis. Image acquisition was performed at a frequency of 3-5 times per week per hospitalized patient to observe temporal changes in the population and size distribution of platelet aggregates under various conditions such as anticoagulant therapy, mechanical ventilation, and discharge from the hospital (Supplementary Table 2). Figure 1d shows a library of typical bright-field images (67 × 67 pixels/image) of single platelets and platelet aggregates flowing at a high speed of 1 m/s, acquired within a field of view of 53.6 μm x 53.6 μm with a spatial resolution of 0.8 μm. As shown in the figure, the ability of our method to directly image and resolve single platelets, platelet-platelet aggregates, platelet-leukocyte aggregates, and other unimportant components (e.g., residual red blood cells, leukocytes, and cell debris) makes it possible to differentiate them with a much higher accuracy than conventional flow cytometry^21,22,23,24^ (especially in discriminating between single platelets and small platelet aggregates). Importantly, this big image database of platelet aggregates enables the previously unfeasible application of advanced computational tools (e.g., high-dimensional analysis and deep learning)^29,40^ which provide unique and insightful analytical results, as we demonstrate here. To the best of our knowledge, this is the first report of image-based profiling and analysis of platelet aggregates on such a statistically large scale, not just for COVID-19, but for any disease.

**Fig. 1.**
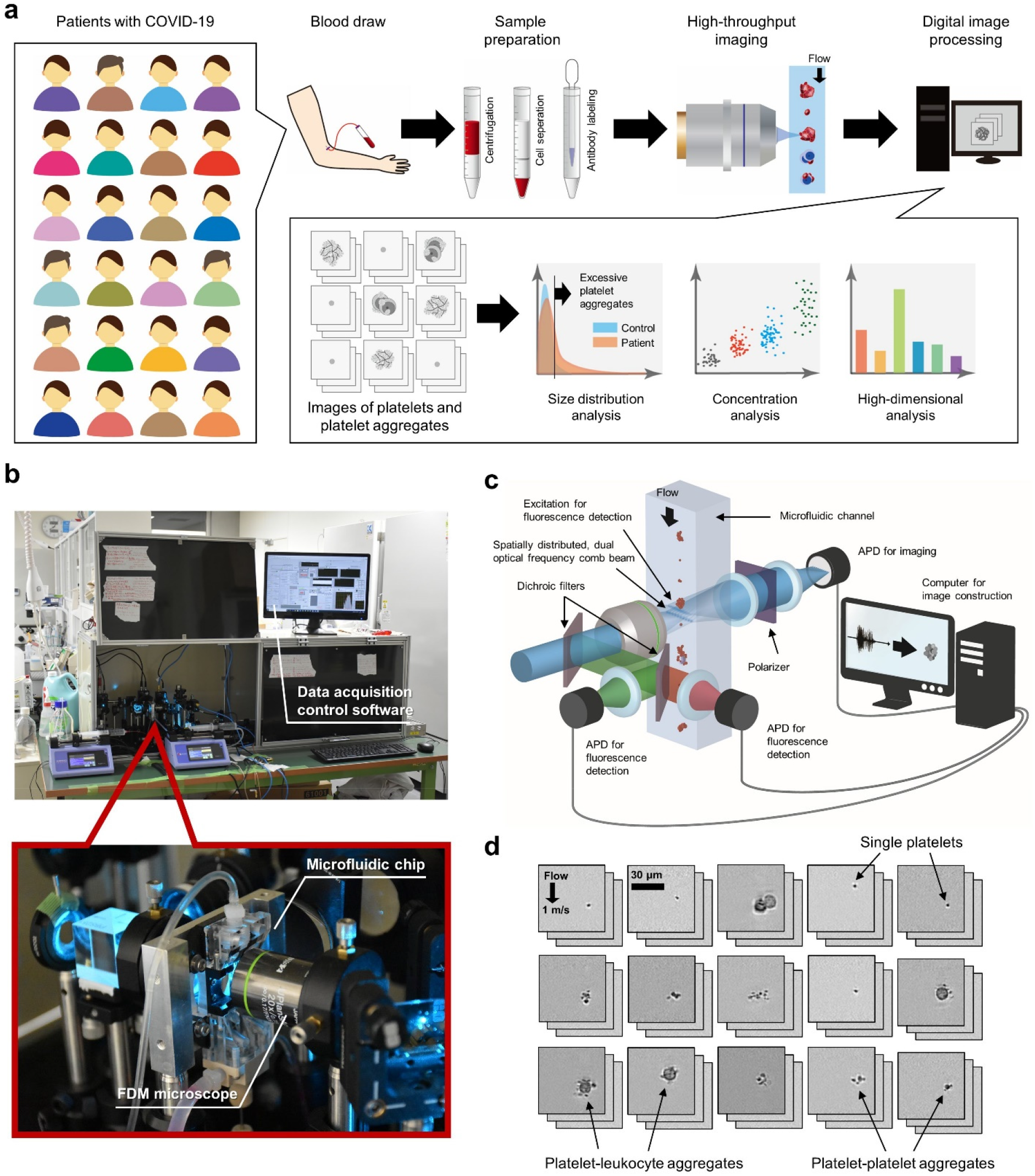
Massive image-based profiling of circulating platelets and platelet aggregates at single-cell resolution. **a**, Experimental workflow consisting of blood draw, sample preparation, high-throughput imaging, and digital image analysis. **b**, Pictures of the FDM microscope on the microfluidic chip installed in the Department of Clinical Laboratory at the University of Tokyo Hospital. The inset shows an enlarged view of the microfluidic chip with the FDM microscope. **c**, Schematic of the FDM microscope for high-throughput, blur-free, bright-field image acquisition. APD: avalanche photodetector. **d**, Typical bright-field images of single platelets and platelet aggregates acquired by the FDM microscope.

## RESULTS

### Image acquisition and size distribution

We acquired 25,000 bright-field images of single platelets and platelet aggregates in the blood of hospitalized patients (n = 110) who were clinically diagnosed with COVID-19 based on their reverse transcription polymerase chain reaction (RT-PCR) test results (Figure 2a, Supplementary Table 1). Negative control image data were also obtained from healthy subjects under the same sample preparation and image acquisition conditions on the same day to mitigate potential bias in the image data that may have come from experimental variations in blood draw, sample preparation, optical alignment, and hydrodynamic focusing conditions (see “sample preparation” in the Methods section for details of the sample preparation protocol) and hence to maintain the state of platelet aggregation in vivo while minimizing the effect of aggregation in vitro. Image acquisition was performed at a high throughput of 100 - 300 events per second (eps), where an event is defined as a single platelet or a platelet aggregate. Residual components such as red blood cells, leukocytes (excluding those contained in platelet-leukocyte aggregates), and cell debris were ignored and not detected as events. The throughput was chosen to avoid clogging the microchannel in the microfluidic chip, although the theoretical throughput of the FDM microscope was >10,000 eps.

**Fig. 2.**
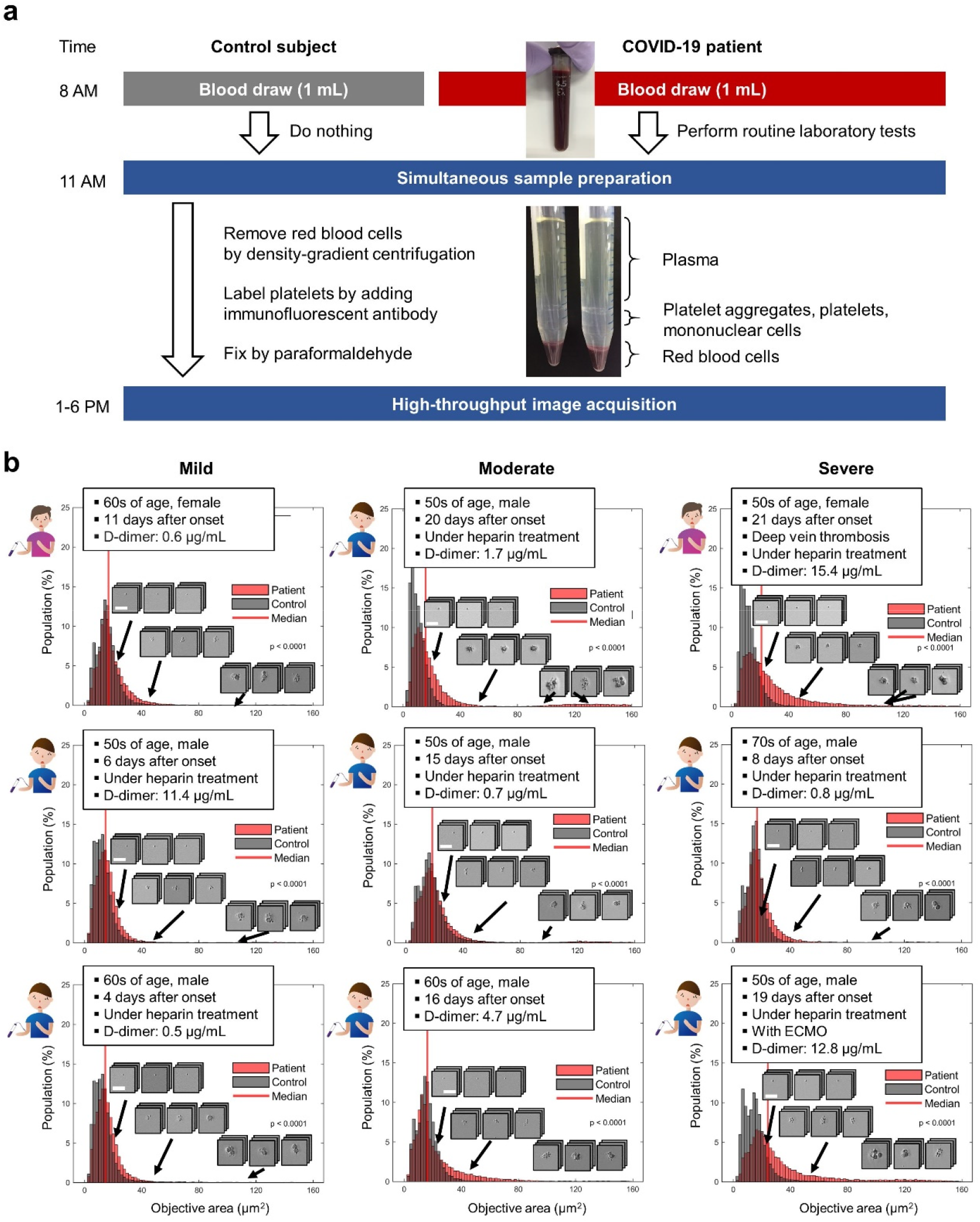
Circulating platelet aggregates in the blood of COVID-19 patients. **a**, Sample preparation protocol. Control data were also obtained under the same sample preparation and image acquisition conditions on the same day to mitigate potential bias in the image data that may have come from experimental variations in blood draw, sample preparation, optical alignment, and hydrodynamic focusing conditions. See “Sample preparation” in the Methods section for details of the protocol. **b**, Size distribution histograms and typical images of single platelets and platelet aggregates in the blood of mild, moderate, and severe patients with COVID-19. The clinical information and D-dimer level of each patient are also shown in the insets. Control data are also shown as references.

Shown in Figure 2b are size distribution histograms and typical images of single platelets and platelet aggregates identified in the blood of patients with COVID-19 (see the “human subjects” in the Methods section and the complete dataset from all 110 patients in Source Data 1). The clinical information and D-dimer level of each patient are also shown in the figure insets. The negative control data are also shown in the figure as references. The objective area was defined by the area of a detected event (i.e., a single platelet or a single platelet aggregate) within each image. The COVID-19 patients were categorized into three groups: (i) the mild patient group: those requiring no oxygen therapy; (ii) the moderate patient group: those requiring oxygen therapy; and (iii) the severe patient group: those requiring mechanical ventilation or extracorporeal membrane oxygenation (ECMO) for respiratory support. Each patient’s highest severity level during his/her hospitalization was used for the categorization. As shown in Figure 2b, excessive platelet aggregates were also present in the blood of patients with low D-dimer levels (≤1 µg/mL), which suggests the presence of enhanced platelet activity by COVID-19, but before the potential onset of clinically evident thrombosis (i.e., the formation and degradation of microthrombi).

### Statistical analysis

We performed a statistical analysis of the big image data of platelet aggregates to visualize trends and correlations. Figure 3a compares the platelet aggregate concentrations of mild, moderate, and severe COVID-19 patients on the highest concentration day of each hospitalized patient. The concentration of platelet aggregates was defined by the ratio of the number of acquired images containing platelet aggregates to the total number of acquired images (n = 25,000) in each sample. Negative control data (shown as “control” in Figure 3a, Figure 3b, and Figure 3c) were provided by blood samples from healthy subjects (n = 4) on 67 different dates. Interestingly, the figure shows the anomalous presence of excessive platelet aggregates in as many as 87.3% of all COVID-19 patients, including those with D-dimer levels below the reference level of ≤1 µg/mL at the University of Tokyo Hospital (see “statistical analysis” in the Methods section for details and see the Discussion section for interpreting the concentration of platelet aggregates in the remaining 12.7% of patients). The excessive platelet aggregation identified in each individual sample is consistent with earlier reports on averaged platelet hyperactivity detected by platelet function tests and gene expression analysis^41,42,43^. The figure also indicates a strong link between the severity of patients and the concentration of platelet aggregates. Moreover, results show an increase in the concentrations of platelet aggregates between measurements taken on the first day after admission (Figure 3b) and on the highest concentration day (Figure 3a) (typically ∼1 week after the first measurement day), in agreement with previous reports on COVID-19 patients whose condition worsened ∼1 week after their initial symptoms appeared^1,5,6,33^. Likewise, Figure 3c shows a strong correlation between the mortality of patients and the concentration of platelet aggregates. Figure 3d, Figure 3e, and Figure 3f show receiver operating characteristic (ROC) curves of the concentration of platelet aggregates with respect to the control corresponding to the data shown in Figure 3a, Figure 3b, and Figure 3c, respectively. The large value of the area under the curve (AUC) in each ROC curve indicates the excellent performance of the concentration of platelet aggregates in evaluating COVID-19-associated platelet activity.

**Fig. 3.**
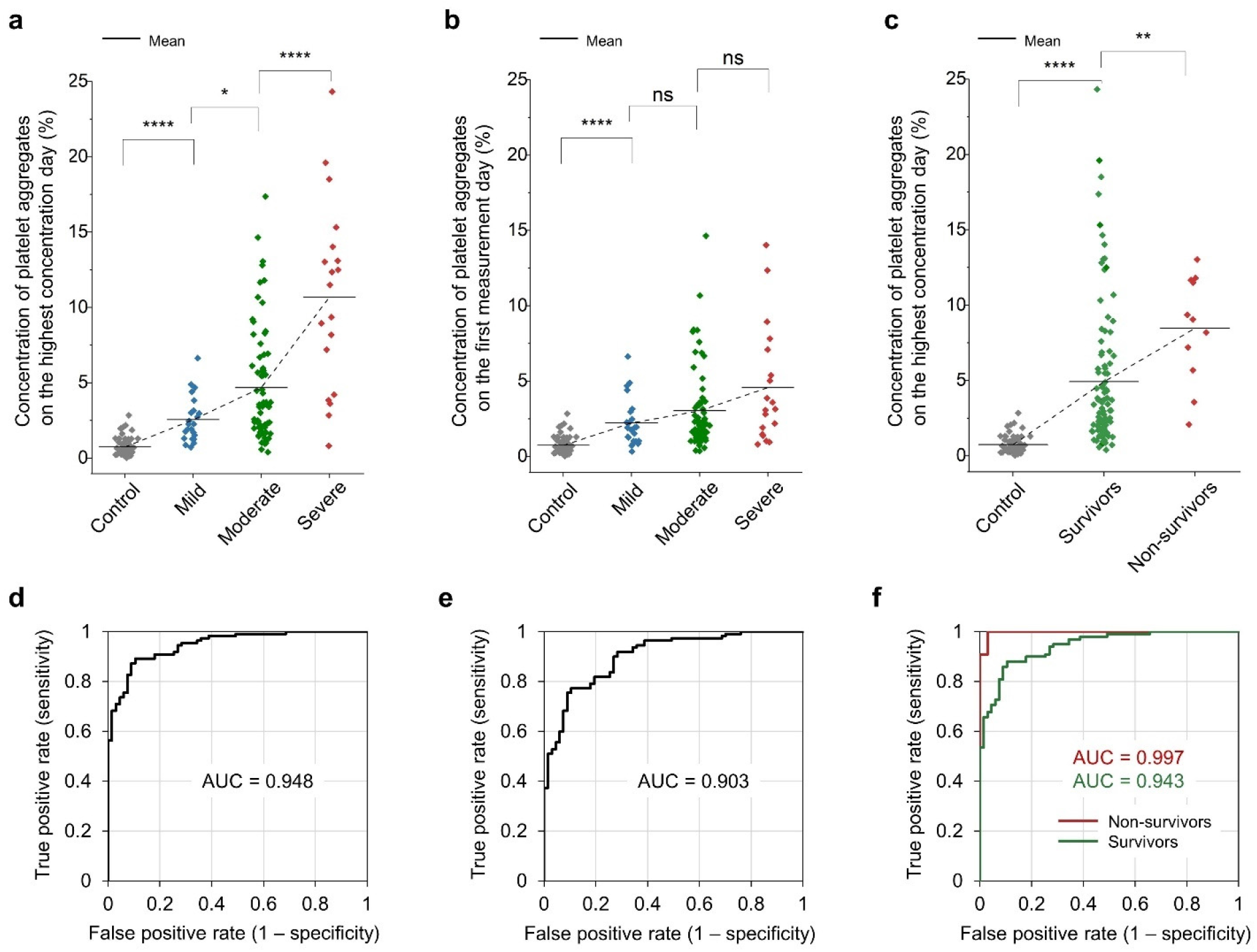
Comparison with the severity and mortality of COVID-19. ns: p > 0.05; *: p ≤ 0.05; **: p ≤ 0.01; ***: p ≤ 0.001; ****: p ≤ 0.0001. **a**, Comparison of the platelet aggregate concentrations of mild (n = 23), moderate (n = 68), and severe (n = 19) patients with COVID-19, measured on the highest concentration day of each hospitalized patient. For the definition of images containing platelet aggregates, see “statistical analysis” in the Methods section for details. **b**, Comparison of the platelet aggregate concentrations of mild (n = 23), moderate (n = 68), and severe (n = 19) patients with COVID-19 on the first measurement day of each hospitalized patient. **c**, Comparison of the platelet aggregate concentrations of survivors (n = 99) and non-survivors (n = 11) from COVID-19, measured on the highest concentration day of each hospitalized patient. **d, e, f**, ROC curves of the data shown in Figure 3a, Figure 3b, and Figure 3c, respectively.

Moreover, we investigated the influence of sex differences and leukocytes. Specifically, Figure 4a shows a higher concentration of platelet aggregates in male patients (n = 73) than in female patients (n = 37) although their statistical significance is not very high, which is aligned with earlier reports that male patients are more prone to intensive treatment unit admission and death than female patients^44^. In addition, the ability of our method to highly resolve platelet aggregates and apply morphometric analysis to their images enabled identifying the presence of leukocytes. Figure 4b and Figure 4c show that the presence of leukocytes in platelet aggregates is linked with the severity and mortality of COVID-19, respectively, which is also consistent with previous reports on leukocyte hyperactivity in COVID-19^26,27,28,33^. Figure 4d and Figure 4e show ROC curves of the concentration of platelet aggregates with respect to the control corresponding to the data shown in Figure 4b and Figure 4c, respectively. The AUCs in these ROC curves indicate the excellent performance of the concentration of platelet aggregates containing leukocytes in evaluating COVID-19-associated platelet-leukocyte activity.

**Fig. 4.**
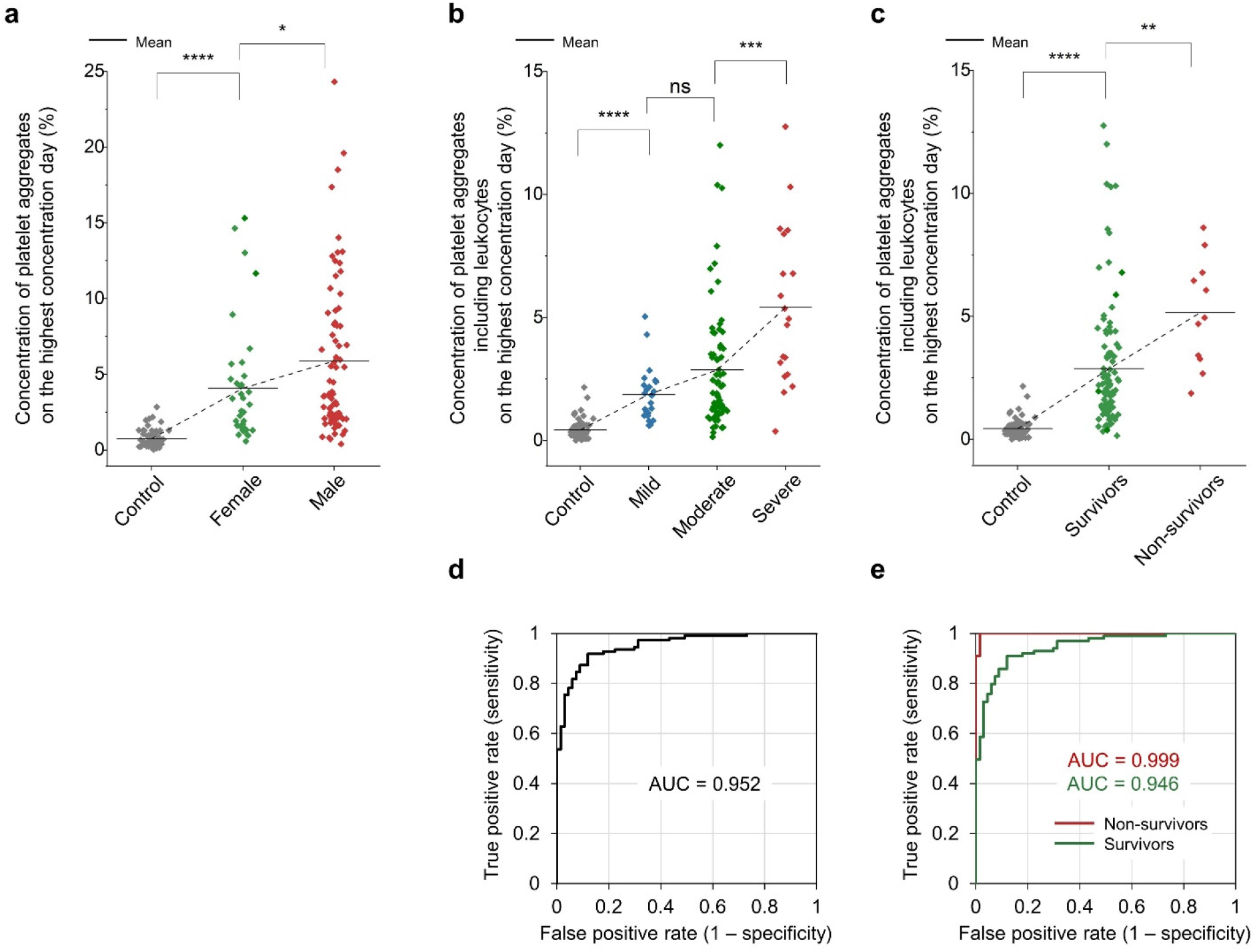
Influence of sex differences and leukocytes. ns: p > 0.05; *: p ≤ 0.05; **: p ≤ 0.01; ***: p ≤ 0.001; ****: p ≤ 0.0001. **a**, Comparison of the platelet aggregate concentrations of male (n = 73) and female (n = 37) patients, measured on the highest concentration day of each hospitalized patient. **b**, Comparison of the concentrations of platelet aggregates of mild (n = 23), moderate (n = 68), and severe (n = 19) patients with COVID-19 including leukocytes, measured on the highest concentration day of each hospitalized patient. **c**, Comparison of the concentrations of platelet aggregates of survivors (n = 99) and non-survivors (n = 11) from COVID-19, measured on the highest concentration day of each hospitalized patient. **d, e**, ROC curves of the data shown in Figure 4b and Figure 4c, respectively.

### Comparison with clinical laboratory and physical findings

We compared our method’s results with those of conventional clinical laboratory and physical findings (Supplementary Table 1, Supplementary Table 3, see “clinical laboratory tests” in the Methods section for details). As shown in Figure 5a, the leukocyte count (WBC), D-dimer level, coagulation factor VIII activity level (FVIII), von Willebrand factor activity level (VWF:RCo), thrombomodulin level (TM), and oxygen administration severity level (respiratory severity) were found to be statistically relevant with the concentration of platelet aggregates measured on the highest concentration day, as indicated by Spearman’s rank correlation coefficients greater than 0.4 for all these parameters (Supplementary Figure 2), whereas the other parameters such as the red blood cell count (RBC), C-reactive protein concentration (CRP), fibrinogen level (Fbg), creatinine concentration (Cre), and body temperature were weakly correlated or uncorrelated with it (Supplementary Figure 3). Additionally, it should be mentioned that the correlation between the concentration of platelet aggregates and the SpO_2_ (oxygen saturation level) level is reasonable since oxygen administration required to maintain the SpO_2_ level in good condition reflects the disease state of patients. These strong correlations between the concentration of platelet aggregates and the WBC, FVIII, VWF:RCo, and TM levels indicate that the high concentration of platelet aggregates is linked with systemic thrombus formation and fibrinolysis, which are expressed by the high D-dimer level, and with vascular endothelial damage (i.e., vasculitis), which is expressed by the high FVIII, VWF:RCo, and TM levels. These links are consistent with earlier reports on severe vascular endothelial damage in the lungs and widespread microthrombi in the alveolar capillaries of COVID-19 patients^3,10,45^.

**Fig. 5.**
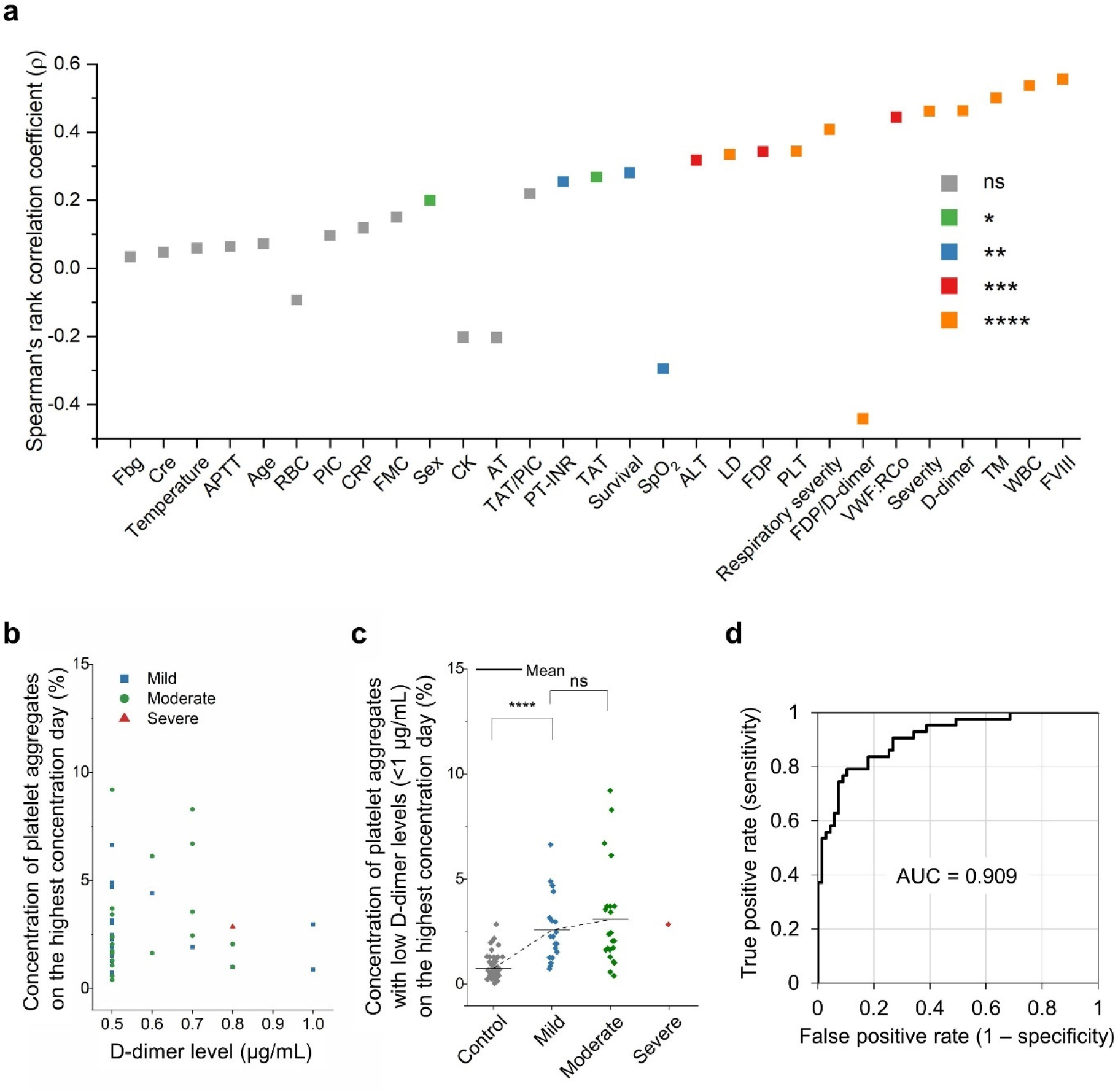
Comparison with clinical laboratory tests. **a**, Comparison of the platelet aggregate concentration measured on the highest concentration day with clinical laboratory and physical findings. WBC: leukocyte count; RBC: red cell count; PLT: platelet count; APTT: activated partial thromboplastin time; PIC: plasma plasmin-α2-plasmin inhibitor complex; CRP: C-reactive protein concentration; FMC: fibrin monomer complex; CK: creatinine kinase; AT: antithrombin; PT-INR: prothrombin time international normalized ratio; TAT: thrombin antithrombin complex; SpO_2_: oxygen saturation; ALT: alanine transaminase concentration; LD: lactate dehydrogenase concentration; FDP: fibrinogen/fibrin degradation product; VWF:RCo: von Willebrand factor activity; TM: thrombomodulin; FVIII: coagulation factor VIII activity; Temperature: body temperature. **b**, Concentrations of platelet aggregates of patients with COVID-19 (39.1% of n = 110) and low D-dimer levels (≤1 µg/mL). **c**, Comparison of the platelet aggregate concentrations of mild (n = 17), moderate (n = 22), and severe (n = 1) patients with COVID-19 and low D-dimer levels (≤1 µg/mL), measured on the highest concentration day of each hospitalized patient. ns: p > 0.05; *: p ≤ 0.05; **: p ≤ 0.01; ***: p ≤ 0.001; ****: p ≤ 0.0001. **d**, ROC curve of the data shown in Figure 5c.

To further investigate significant explanatory factors for the concentration of platelet aggregates, we performed a multivariate regression analysis (SPSS software, IBM) using the vascular endothelial disorder marker group (FVIII, TM, VWF:RCo), the coagulation / fibrinolysis marker group (D-dimer, FDP, TAT, PT-INR), and the group of other markers having strong correlations with the concentration of platelet aggregates on the highest concentration day (WBC, respiratory severity, PLT, LD, ALT, SpO_2_, survival, sex) as potential explanatory factors (Supplementary Table 4). As a result, the analysis identified FVIII, PLT, WBC, PT-INR, and respiratory severity as significant predictors of the concentration of platelet aggregates on the highest concentration day. A further multivariate regression analysis using these parameters identified respiratory severity (p = 0.009) and FVIII (p = 0.016) as the most significant predictors of the concentration of platelet aggregates on the highest concentration day.

Next, since earlier reports show the onset of COVID-19-associated thrombosis even with low D-dimer levels^4^, we investigated patient cases with D-dimer levels below the reference level of ≤1 µg/mL, which constitutes 39.1% of all COVID-19 patients (n = 110). As shown in Figure 5b, significant levels of platelet aggregate concentrations are evident. Also, as shown in Figure 5c, the correlation between the severity of patients and the concentration of platelet aggregates also persists and is similar to the relation shown in Figure 3a although the difference between the mild and moderate patients is not very large, which is reasonable because their medical conditions (both without mechanical ventilation or ECMO) are similar. The figure also shows that excessive platelet aggregates were found in as many as 76.7% of patients with a D-dimer level of ≤1 µg/mL (see “statistical analysis” in the Methods section for details). These results may suggest the presence of a non-negligible microvascular thrombotic risk that could not be detected by the D-dimer test. In other words, the D-dimer test mainly evaluates blood coagulation by measuring D-dimers that are produced either at the final stage of thrombus formation (i.e., cross-linking) or after thrombus degradation^32^ and is known to be insensitive to microvascular thrombosis including TMA^35^, whereas our method characterizes the initiation of microthrombus formation and the degradation of microthrombi^15,16,17,18^. The latter method is, therefore, more sensitive with high resolution to platelet hyperactivity, which is a suggested mechanism of widespread microthrombus formation^2,41^. In addition, the fact that only one severe case with a low D-dimer level was identified as shown in Figure 5c can be explained by recognizing that the D-dimer test is generally insensitive to TMA as mentioned above, but can detect it if the medical conditions of patients become severe^46^. In other words, the correlation between the D-dimer level and the concentration of platelet aggregates in Figure 5a is valid for moderate-severe cases while not significant for mild-moderate cases. Finally, as shown in Figure 5d, the large AUC value of the ROC curve of the concentration of platelet aggregates with low D-dimer levels of ≤1 µg/mL with respect to the control indicates our method’s high performance in assessing the severity of COVID-19 even when the D-dimer level is low.

### Temporal monitoring

Our temporal monitoring of the concentration of platelet aggregates in the blood of COVID-19 patients enabled us to probe the pathological conditions of COVID-19 patients, most of whom received anticoagulant therapy with heparin. Figure 6a shows the evolution of the platelet aggregate concentration of each patient group (mild, moderate, severe) after the onset of COVID-19 (see “human subjects” in the Methods section, the complete dataset in Source Data 1, and the measurement days in Supplementary Table 2). As mentioned above, each patient’s highest severity level during his/her hospitalization was used for the severity categorization. Specifically, as shown in the figure, the platelet aggregate concentration of the mild patient group reached a peak (about 2.5%) in the first 9 – 12 days and then gradually decreased over a week, followed by the complete discharge of all the mild patients from the hospital after 16 days. Likewise, the platelet aggregate concentration of the moderate patient group reached a peak (about 5%) after the first 13 – 16 days and then gradually decreased over two weeks, followed by the complete discharge of all the moderate patients from the hospital after 28 days.

**Fig. 6.**
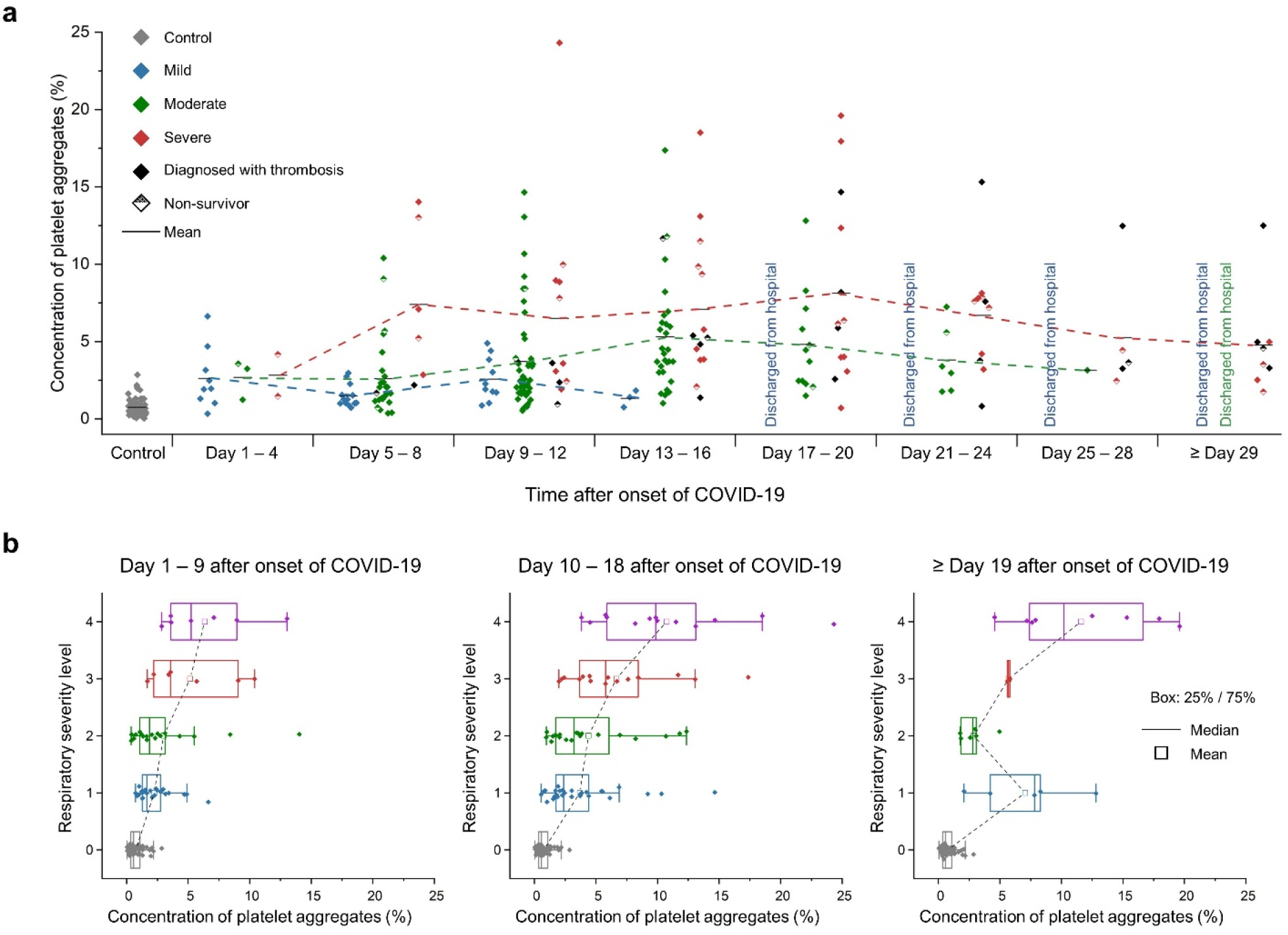
Temporal monitoring. **a**, Evolution of the platelet aggregate concentrations of different severity groups (mild, moderate, severe). Each patient’s highest severity level during his/her hospitalization was used for the severity categorization and fixed during the entire hospitalization period to trace the evolution of the platelet aggregation concentration of each patient group. **b**, Comparison of the respiratory severity level with the concentration of platelet aggregates. Level 0: control. Level 1: without oxygen administration; Level 2: with oxygen administration of 0.5 – 4 L/min; Level 3: with oxygen administration of ≥ 5 L/min; Level 4: with mechanical ventilation or ECMO.

On the other hand, the platelet aggregate concentration of the severe patient group initially increased to a peak (about 7%) in the first week and then exhibited a plateau for three weeks, followed by death or transfer to a chronic hospital. It is important to note that all the patient groups showed a moderately high platelet aggregate concentration level (about 2.5%) in the first 3 – 4 days, but their concentrations started to differ in the next 3 – 4 days, such that each patient group underwent a different prognosis pattern while the timing of discharge from the hospital was consistent with the decreased concentration of platelet aggregates in all prognosis patterns. Also, as shown in the figure, patients who developed thrombosis ended up staying in the hospital longer than the mild and moderate patients and some of the severe patients. Moreover, as shown in Figure 6b, the strong correlation between the respiratory severity level and the concentration of platelet aggregates in the first two periods (day 1 – 9, day 10 – 18) indicates that the concentration of platelet aggregates is a good indicator of the respiratory condition of COVID-19 patients. The twin-peak distribution of the platelet aggregate concentrations of patients in the last period (≥ day 19 after the onset of COVID-19) indicates that most patients either ended up recovering or remained hospitalized as characterized by the patients at levels 2 and 3 (those requiring oxygen administration) stepping down to level 1 (no oxygen administration) and discharging from the hospital or the patients staying at level 4 (requiring mechanical ventilation or ECMO), respectively.

### Comparison with other diseases

We compared the concentrations of platelet aggregates from patients with COVID-19 and patients with other diseases that are known to produce platelet aggregates (Figure 7a, Supplementary Table 5, Supplementary Figure 4, see “high-dimensional analysis” in the Methods section for details). Interestingly, the average concentration of platelet aggregates in COVID-19 is higher than that in thrombosis (e.g., stroke, deep vein thrombosis, pulmonary embolism; n = 16) and is close to that in other infectious diseases (e.g., pneumonia, pyelonephritis, cholangitis; n = 7) which are known to cause cytokine-mediated coagulation activation and the formation of neutrophil extracellular traps. The results can be understood by recognizing that COVID-19 is a systemic disease caused by microvascular thrombosis via persistent endothelial cell activation upon SARS-CoV-2’s infection of the vascular endothelium^2,47^, resulting in platelet aggregation which may or may not involve coagulation characterized by D-dimer production^1,48^, whereas thrombosis is a localized disease in which the platelet aggregates produced at the site of thrombi are diluted in the circulation. Additionally, as shown in Figure 7b, our deep-learning-based high-dimensional comparison of the images of platelet aggregates in COVID-19 with those in thrombosis and other infectious diseases, using the same image data shown in Figure 7a, shows that while platelet aggregates in non-COVID-19 infectious diseases and non-COVID-19 thrombosis are clearly different, platelet aggregates in COVID-19 have properties of both disease types. Likewise, as shown in Figure 7c, our comparison of the images of platelet aggregates in COVID-19 without risk factors for thrombosis (i.e., without thrombosis-related underlying conditions) (n = 22) with those in non-COVID-19 with risk factors only for venous thrombosis (cancer, post-surgery, long-term bed rest, obesity, and heart failure; n = 18) and with risk factors only for arterial thrombosis (hypertension, diabetes, hyperlipidemia, smoking, and history of atherosclerosis; n = 15) indicates that while platelet aggregates in the two risk factor groups are distinct, platelet aggregates in COVID-19 have characteristics of both groups, with a stronger overlap with the venous thrombosis risk factor group, which is consistent with earlier reports on the diversity of COVID-19-associated thrombosis^1^ as well as the statistics of its types as reported by international societies on thrombosis and hemostasis (arterial thrombosis: ∼30%; venous thrombosis: ∼70%). Although thrombosis and infectious diseases are known discretely as localized and systemic diseases, respectively, our results indicate that COVID-19 cannot be classified easily into either category; COVID-19 instead behaves as a product of the two disease classes, as a cause of systemic thrombosis, which agrees with the widely accepted mechanism of COVID-19^2^.

**Fig. 7.**
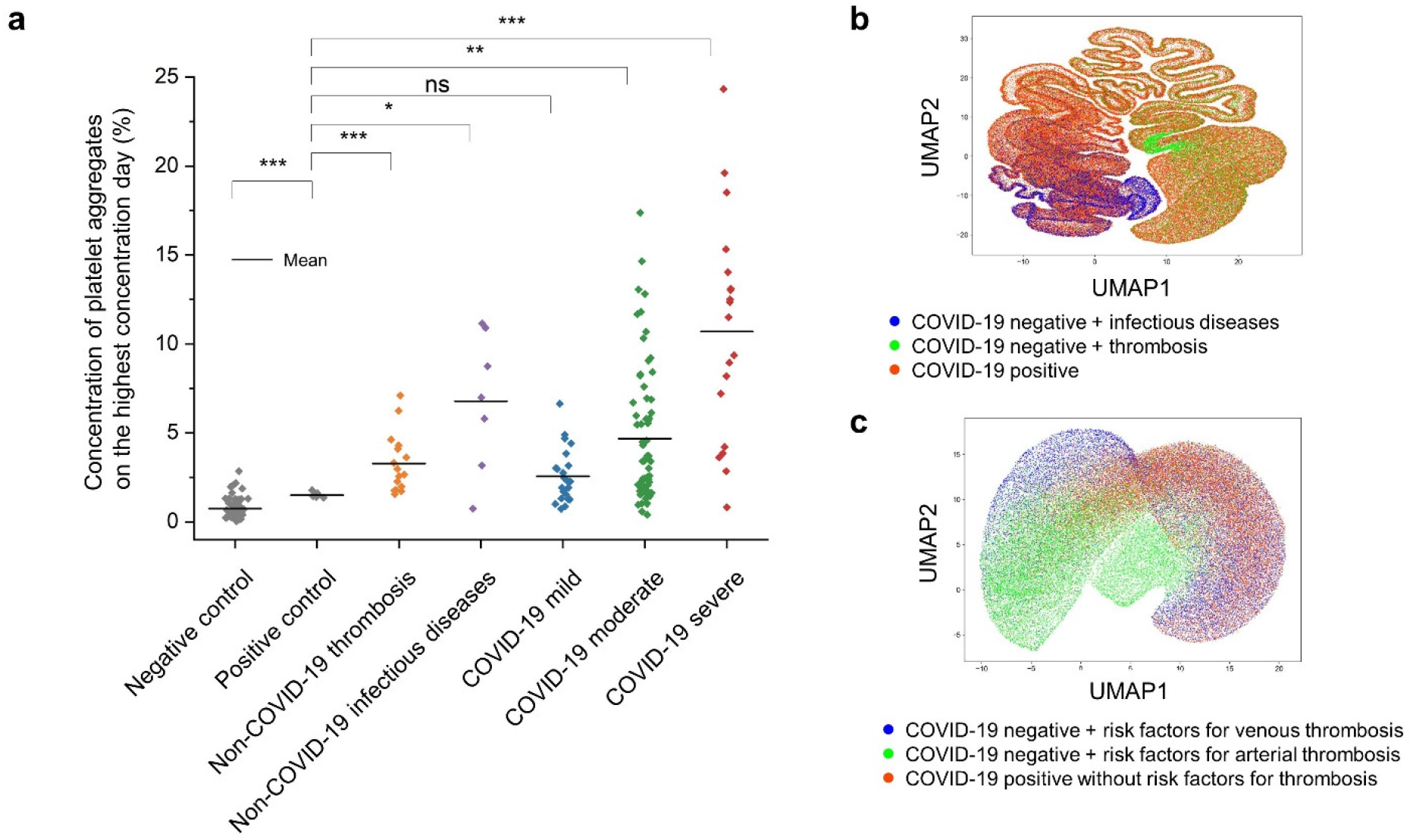
Comparison with other diseases. **a**, Concentrations of platelet aggregates of patients with COVID-19 compared with those of the positive control group and patients with non-COVID-19 thrombosis (n = 16) and non-COVID-19 infectious diseases (n = 7). Negative control: healthy subjects; positive control: patients with diseases that do not produce platelet aggregates; thrombosis: patients with stroke, deep vein thrombosis, pulmonary embolism, etc. Non-COVID-19 infectious diseases below the disseminated intravascular coagulation (DIC) diagnostic criteria: patients with pneumonia, pyelonephritis, cholangitis, etc. ns: p > 0.05; *: p ≤ 0.05; **: p ≤ 0.01; ***: p ≤ 0.001; ****: p ≤ 0.0001. **b**, UMAP plot of platelet aggregates from COVID-19 patients, non-COVID-19 infectious diseases, and non-COVID-19 thrombosis, using the same data shown in Figure 7a. **c**, UMAP plot of platelet aggregates from COVID-19 patients without risk factors for thrombosis (n = 22), non-COVID-19 patients with risk factors for venous thrombosis (n = 18), and non-COVID-19 patients with risk factors for arterial thrombosis (n = 15).

## DISCUSSION

By virtue of its combined capability of massive image-based profiling, temporal monitoring, and big data analysis of circulating platelets and platelet aggregates in the blood of COVID-19 patients, our method is a powerful tool for offering new insights into the underlying process of COVID-19-associated microvascular thrombosis. Specifically, it has shown the strong link between the concentration of platelet aggregates and the severity and mortality of COVID-19. Furthermore, our high-dimensional analysis of platelet aggregate images and comparison with images of platelet aggregates in other diseases revealed that COVID-19 behaves as a systemic thrombosis, which suggests the coordinated activation of the inflammatory and thrombotic responses (i.e., thromboinflammation) to SARS-CoV-2 infection via vascular endothelial damage. This is aligned with our current understanding of the SARS-CoV-2 infection mechanism^6^ and is, in fact, strongly supported by the positive correlation between the concentration of platelet aggregates and the endothelial activation characterized by the VWF:RCo, FVIII, and TM levels (Figure 5a). Specifically, in the widely accepted pathophysiological mechanism of COVID-19, the viral entry into host cells is mediated by the ACE2 receptor, causing vascular endothelial damage and resulting in vasculitis (i.e., the inflammation of blood vessels)^2,6^. Since VWF is a factor that mediates platelet adhesion and aggregation at the site of vascular endothelial injury, it is released into the blood in large amounts during the inflammation-mediated endothelial injury. Our findings (Figure 3a, Figure 5a) verified earlier reports on the correlation between the VWF level and the severity of COVID-19^47^ by directly observing the increased concentration of platelet aggregates with its positive link to the severity of COVID-19. Additionally, our findings (Figure 3a through Figure 3c) also support a recently suggested mechanism in which platelets are hyperactive during COVID-19, not just via vasculitis, but also via the direct interaction of SARS-CoV-2 with platelets^43^, which may account for the anomalous presence of excessive platelet aggregates found in nearly 90% of all COVID-19 patients in our study. Finally, based on the earlier reports that the lung is a major site of platelet biogenesis^49^ and develops widespread microthrombi in severe COVID-19 patients^3^, it can be speculated that the simultaneous activation of platelets and endothelial cells in the lung may be closely involved in the deterioration of respiratory conditions in COVID-19 patients and antiplatelet therapy may be effective for arterial oxygenation and improving clinical outcomes in severe COVID-19 patients.

Our findings suggest that measuring the concentration and distribution of circulating platelet aggregates is a potentially effective approach to evaluating the potential risk of microthrombus formation. In fact, there are a number of postmortem examination reports showing that the primary cause of death in patients who died of COVID-19 pneumonia was respiratory failure due to diffuse alveolar damage with severe capillary congestion caused by microthrombi^10,16^. The strong correlation between the concentration of platelet aggregates and the oxygen administration severity and SpO_2_ levels (Figure 5a, Figure 6a, Figure 6b) strongly suggests our method’s ability to detect precursors to widespread microthrombus formation. Also, in our study, COVID-19 patients who were not diagnosed with thrombosis may have developed microthrombi which were too small to detect by medical imaging (e.g., CT and MRI), but were identified by our method as shown in Figure 3a and Figure 3c. In fact, a meta-analysis of COVID-19-associated thrombosis has reported that the incidence of venous thromboembolism differed significantly, depending on whether a screening test was performed or not^50^. It is possible that the incidence of thrombosis was missed in our clinical diagnosis, which may account for the mismatch between the severe cases and the confirmed onset of thrombosis in Figure 6a. Finally, Figure 5c indicates that our method is sensitive to platelet activity as a potential precursor of microthrombus formation in COVID-19-associated microvascular thrombosis, while the D-dimer test is insensitive to it unless the severity of the patient’s medical condition is high. Further studies are needed to directly verify the link between the concentration of platelet aggregates and the risk of microvascular thrombosis, for example, via comparing results from our method and autopsies.

The reason why excessive platelet aggregates were not found in the remaining 12.7% of the COVID-19 patients can be explained by recognizing two factors. First, the frequency of measurements on the mild and moderate patients was much lower than that of the severe patients, which led to a lower accuracy in finding the optimal timing for measuring the concentration of platelet aggregates (i.e., the highest concentration day) for the mild and moderate patients. In fact, the average numbers of measurements per patient were 1.9 (mild), 2.6 (moderate), and 7.3 (severe) times. The fact that the high AUC in the ROC curve of the concentration of platelet aggregates on the first measurement day is as large as 0.9 (as shown in Figure 3e) indicates that both the sensitivity and specificity of our method are high despite the one-time measurements. This means that the low frequency of measurements is responsible for the lower concentration of platelet aggregates measured. Second, nearly all the COVID-19 patients received heparin-based anticoagulant treatment, which may have led to an overall reduction in the concentration of platelet aggregates. Therefore, more frequent measurements without the influence of anticoagulant therapy (e.g., using animal models) are expected to improve the temporal resolution of image-based platelet aggregate profiling in assessing the varying severity of COVID-19 patients and hence improve the accuracy of the plots, trends, and correlations in Figure 3a, Figure 3c, Figure 4a, Figure 4b, Figure 4c, Figure 5b, and Figure 5c.

There are a few limitations to this study and potential solutions to them. First, all the subjects were hospitalized patients at the University of Tokyo Hospital alone, which may have introduced a selection bias. Further multi-hospital studies are needed to draw more general conclusions. Second, since our analysis is a retrospective observational study, further prospective studies are needed to determine whether the concentration of platelet aggregates can predict worsening respiratory status and the development of microthrombi. With these improvements, our method may hold promise for predicting respiratory failure at an early stage and for developing treatment strategies for preventing the transition of patients to ventilatory management.

## METHODS

### Human subjects

The subjects used in this study were patients who were clinically diagnosed with COVID-19 based on their reverse transcription polymerase chain reaction (RT-PCR) test results (Supplementary Table 1). Blood samples were collected as residual coagulation test samples (with 3.2% citrate) after the completion of requested clinical laboratory tests at the University of Tokyo Hospital. Blood cells in the samples were stored at room temperature while the residual plasma was cryopreserved at -80 °C. The negative control group was composed of 4 healthy subjects. Blood samples (with 3.2% citrate) from the healthy subjects were drawn multiple times on 67 different dates for preparing control samples. Likewise, to verify that the effect of using the residual coagulation test samples after the clinical laboratory tests on platelet aggregation was negligible, the positive control group was composed of 7 hospitalized patients under no anticoagulant therapy and with no abnormality confirmed by their coagulation tests, which indicated prothrombin time international normalized ratio (PT-INR), activated partial thromboplastin time (APTT), and fibrinogen levels of 0.88 - 1.10, 24 - 34 sec, and 1.68 - 3.55 g/L, respectively, and D-dimer levels of less than 1 µg/mL. Subjects under anticoagulant therapy were excluded. Clinical information (e.g., sex, age, severity, SpO_2_ level, body temperature, respiratory severity) and laboratory test data were obtained from the electronic medical records of the patients using a standardized data collection form. For comparison, blood samples (with 3.2% citrate) from subjects with other diseases that elevate D-dimer levels, such as cancer, thrombosis, and sepsis were also analyzed. This study was conducted with the approval of the Institutional Ethics Committee in the School of Medicine at the University of Tokyo [no. 11049, no. 11344] in compliance with the relevant guidelines and regulations. Written informed consent for participation in the study was obtained from the patients using an opt-out process on the webpage of the University of Tokyo Hospital. Patients who refused participation in our study were excluded. Written informed consent was obtained from the healthy subjects as well. The demographics, clinical characteristics, and laboratory findings of patients with COVID-19 are shown in Supplementary Table 1. The demographics, clinical characteristics, and laboratory findings of patients with other diseases (e.g., cancer, non-COVID-19 thrombosis, non-COVID-19 infectious diseases) are shown in Supplementary Table 5.

### Sample preparation

Single platelets and platelet aggregates were enriched from whole blood by density-gradient centrifugation to maximize the efficiency of detecting platelets and platelet aggregates, as described in our previous report^24^ with minor modifications. As shown in Figure 2a, for analyzing the concentration of platelet aggregates, 500 µL of blood was diluted with 5 mL of saline. Platelets were isolated by using Lymphoprep (STEMCELLS, ST07851), a density-gradient medium, based on the protocol provided by the vendor. Specifically, the diluted blood was added to the Lymphoprep medium and centrifuged at 800 g for 20 min. After the centrifugation, 500 µL of the sample was taken from the mononuclear layer. Platelets were immunofluorescently labeled by adding 10 µL of anti-CD61-PE (Beckman Coulter, IM3605) and 5 µL of anti-CD45-PC7 (Beckman Coulter, IM3548) to the blood sample to ensure the detection of all platelets or platelet aggregates in the sample. Then, 500 µL of 2% paraformaldehyde (Wako, 163-20145) was added for fixation. Without the fixation, platelet aggregates in the sample would be dismantled as reported in our earlier work^24^. Therefore, the fixation process was performed at least within 4 hours after the blood draw. The entire sample preparation was performed at room temperature.

### Clinical laboratory tests

Blood samples from the patients were used for routine laboratory tests such as the leukocyte count (WBC), red cell count (RBC), platelet count (PLT), prothrombin time international normalized ratio (PT-INR), activated partial thromboplastin time (APTT), D-dimer level, fibrinogen level (Fbg), C-reactive protein concentration level (CRP), alanine transaminase concentration level (ALT), lactate dehydrogenase concentration level (LD), and creatinine kinase level (CK). The cryopreserved plasma was used for additional tests such as plasma plasmin-α2-plasmin inhibitor complex level (PIC), fibrin monomer complex level (FMC), antithrombin level (AT), thrombin antithrombin complex level (TAT), von Willebrand factor activity level (VWF:RCo), fibrinogen/fibrin degradation product level (FDP), thrombomodulin level (TM), and coagulation factor VIII activity level (FVIII) (Figure 5a, Supplementary Table 1, Supplementary Table 3). All the coagulation tests (i.e., AT, FDP, TAT, FMC, PIC, FVIII, VWF:RCo, TM, and D-dimer) were conducted on a CN6500 automatic coagulation analyzer (Sysmex, Japan). The D-dimer tests were performed with a latex-enhanced photometric immunoassay (LIAS AUTO D-dimer NEO, Sysmex, Japan). The laboratory reference range of the D-dimer test at the University of Tokyo Hospital was 0 - 1.0 µg/mL. Since the FVIII and VWF:RCo values of many blood samples exceeded the upper measurement limits of the analyzer (FVIII: 480%; VWF:RCo: 300%), their values were calculated based on the measured absorbance (FVIII: y = 1090.5x^2^ + 414.52x + 30.776; VWF:RCo: y = 1716x + 30.149; where x = absorbance, y is expressed in units of %).

### Optical frequency-division-multiplexed microscope

The FDM microscope is a high-speed, blur-free, bright-field imaging system based on a spatially distributed optical frequency comb as the optical source and a single-pixel photodetector as the image sensor. Since the optical frequency comb is composed of multiple beams which are spatially distributed, it is capable of simultaneously interrogating the one-dimensional spatial profile of a target object (e.g., a platelet, a platelet aggregate). In addition, since each discrete beam of the optical frequency comb is tagged by a different modulation frequency, a spatial-profile-encoded image can be retrieved by performing Fourier transformation on the time-domain waveform detected by the single-pixel detector. As shown in Supplementary Figure 1, we used a continuous-wave laser (Cobolt Calypso, 491 nm, 100 mW) as the laser source. Emitted light from the laser was split by a beam splitter, deflected and frequency-shifted by acousto-optic deflectors (Brimrose TED-150-100-488, 100-MHz bandwidth), and recombined by another beam splitter. The resultant optical frequency comb was focused by an objective lens (Olympus UPLSAPO20X, NA:0.75) onto objects (e.g., single platelets, platelet aggregates) flowing at 1 m/s in a customized hydrodynamic-focusing microfluidic channel (Hamamatsu Photonics). Light transmitted through the flowing objects was collected by an avalanche photodiode (Thorlabs APD430A/M) and processed by a home-made LabVIEW program to reconstruct the bright-field images. The line scan rate, spatial resolution, field of view, and number of pixels after making pixel aspect ratio corrections were 3 MHz, 0.8 μm, 53.6 μm x 53.6 μm, and 67 × 67 pixels, respectively. Fluorescence emitted from platelets labeled by anti-CD61-PE was also collected and used for triggering image acquisitions. Fluorescence emitted from leukocytes labeled by anti-CD45-PC7 was collected to identify platelet aggregates containing leukocytes. Image acquisition was performed at a high throughput of 100 - 300 events per sec (eps), where an event is defined as a single platelet or a platelet aggregate since red blood cells, leukocytes, and cell debris were not detected as events. The throughput was chosen to avoid clogging the microchannel, although the theoretical throughput of the machine was >10,000 eps.

### Statistical analysis

The regions of objects (i.e., platelets, platelet aggregates) in bright-field images were segmented in MATLAB R2020a for calculating the concentration of platelet aggregates in each sample. First, a 10x interpolation by the interp2 function (MATLAB R2020a) was applied to each image for achieving segmentation results. Then, the outlines of the object regions were detected by using the edge detection function with the Canny method in MATLAB R2020a. Morphological operations like dilate, fill, and erode were applied to fill and refine the object regions for obtaining their masks as well as for eliminating the background noise. After the segmentation, the size of the object (i.e., a single platelet, a platelet aggregate) in each image was calculated by multiplying the pixel size of the segmented region, which was extracted by the regionprops function, and the pixel resolution after interpolation (80 nm x 80 nm/pixel). All the images with objective areas larger than 48 µm^2^ were considered as the images of platelet aggregates in all the patient and healthy subject (control) samples. The concentration of platelet aggregates was defined by the ratio of the number of acquired images containing platelet aggregates to the total number of acquired images (n = 25,000) in each sample. In all 110 patient datasets, 106 of them have 25,000 images while only 4 of them (no. 6, 10, 12, 14) have 20,000 images due to a data-recording error, but this should not influence the statistical accuracy of our data analysis since the number of acquired images is significantly large. The presence of leukocytes in platelet aggregates was also identified by analyzing the morphology of the platelet aggregates by their images. For confirmation, the fluorescence signal of anti-CD61-PE and anti-CD45-PC7 was also used. CD61-PE/CD45-PC7 double-positive events were counted as platelet aggregates containing leukocytes. CD61-PE-positive and CD45-PC7-negative events that had CD61-PE signal intensity greater than a threshold value were counted as platelet aggregates excluding leukocytes. The presence of excessive platelet aggregates was determined by calculating the mean and standard deviation of the distribution of the concentration of platelet aggregates in the control samples and evaluating if the concentration of platelet aggregates in a patient sample exceeded the threshold (mean + standard deviation). If a tighter threshold (mean + two standard deviations) was used to calculate the presence of excessive platelet aggregates, then the ratio of the number of patients with excessive platelet aggregates to the number of all patients is 75.5%, while the ratio of the number of patients with excessive platelet aggregates and low D-dimer levels (≤1 µg/mL) to the number of all patients with low D-dimer levels (≤1 µg/mL) is 62.8%. Statistical analysis was performed using the rank sum test including the Kruskal-Wallis test, Mann–Whitney U test, and Wilcoxon signed-rank test with significance levels at 0.05. Each statistical correlation value was obtained by calculating Spearman’s rank correlation coefficient between the concentration of platelet aggregates on the highest concentration date and the corresponding clinical parameters. The line fitting function of the Origin 2021b software was used to draw linear fits to show the correlation between the concentration of platelet aggregates and each clinical parameter. A 95% confidence interval is shown as a gray area in each panel (Supplementary Figure 2, Supplementary Figure 3).

### High-dimensional analysis

In both Figure 7b and Figure 7c, a fully connected classifier was trained on image data from non-COVID-19 patient blood samples under specific classes to build a visual feature extractor. The class label was represented as a one-hot vector. Images were picked with size gating and normalized to zero mean and unit variance. The images were input into VGG-16 with pre-trained weights on ImageNet dataset. The output of the fourth pooling layer was flattened into an 8192-dimensional vector and input into the classifier. The classifier was optimized by Adam optimizer with loss function of mean absolute error. To improve classification accuracy for under-represented classes, the loss function was weighted by inverse class frequency. The learning rate was reduced from 0.01 by a factor of 0.31 when the validation loss stopped decreasing for more than 10 epochs. The training was terminated when there was no significant improvement for 100 epochs. The training and validation set was randomly selected with an 80/20 ratio. After the training, the final layer was removed and the rest was used as a feature extractor, which output a 48-dimensional vector. The dimension of feature vectors was reduced into minimum dimension by principal component analysis, so that the cumulative contribution rate of the selected principal components was more than 0.99. For Figure 7b, 13770 images of platelet aggregates from 16 thrombosis patients and 12651 images of platelet aggregates from 7 infectious disease patients were used for the training and validation and displayed in the uniform manifold approximation and projection (UMAP) plot. 150116 images of platelet aggregates from 110 COVID-19 patients were added to the UMAP plot. The feature extractor was implemented in Tensorflow. VGG-16 was used as the application in the library. Figure 7b was plotted with parameters n_neighbor = 1000 and min_dist = 1. For Figure 7c, 19649 images of platelet aggregates from 15 patients with risk factors for arterial thrombosis (hypertension, diabetes, hyperlipidemia, smoking, and history of atherosclerosis) and 20850 images of platelet aggregates from 18 patients with risk factors for venous thrombosis (cancer, post-surgery, long-term bed rest, obesity, and heart failure) were used for the training and validation and displayed in the UMAP plot. Patients in each group did not have risk factors for the diseases in the other groups. To show their relations with the diseases, 22890 images of platelet aggregates from 22 COVID-19 patients without thrombosis-related underlying conditions were added to the UMAP plot. Figure 7c was plotted with n_neighbor = 4000 and min_dist = 1.

## Data Availability

The source data (Source Data 1) supporting the findings of this study are available from the corresponding authors upon reasonable request.

## Data availability

The source data (Source Data 1) supporting the findings of this study are available at http://doi.org/10.5281/zenodo.4700112 and are also available from the corresponding authors upon reasonable request.

## Code availability

All the code used for data taking and analysis in this study is available at http://doi.org/10.5281/zenodo.4700072 and is also available from the corresponding authors upon reasonable request.

## Acknowledgements

This work was supported by AMED JP20wm0325021, JSPS Core-to-Core Program, JSPS KAKENHI grant numbers 19H05633 and 20H00317, ImPACT Program (CSTI, Cabinet Office, Government of Japan), White Rock Foundation, Ogasawara Foundation, Nakatani Foundation, Konica Minolta Foundation, and Charitable Trust Laboratory Medicine Research Foundation of Japan. We thank Kyoko Hasegawa and Yoshika Kusumoto for help with the sample preparation.

## Author information

### Contributions

K. G. conceived the work. H. K., Y. Z., T. X., and K. G. designed the machine for massive image-based profiling of platelet aggregates. H. K. built the machine. M. N., J. T., and Y. Z. prepared the blood samples. H. K., Y. I., and M. N. performed the image acquisition of platelets and platelet aggregates. Y. Z., S. T., M. R., T. S., and G. R. analyzed the image data. J. H., S. T., Y. D., H. Z., K. H., R. K., W. P., M. S., Y. Z. X. Y., A. R., G. R., and W. I. helped the image data acquisition and analysis. Y. Z., M. N., T. X., N. N., S. M., Y. Y., and K. G. interpreted the analysis results. T. X., Y. Y., and K. G. supervised the work. All others participated in writing the paper.

## Ethics declarations

N. N. and K. G. are shareholders of CYBO, Inc. The other authors have no competing interests.

